# Multicenter Evaluation of Interpretable AI for Coronary Artery Disease Diagnosis from PET Biomarkers

**DOI:** 10.1101/2025.06.19.25329944

**Authors:** Wenhao Zhang, Jacek Kwiecinski, Aakash Shanbhag, Robert JH. Miller, Giselle Ramirez, Jirong Yi, Donghee Han, Damini Dey, Dominika Grodecka, Kajetan Grodecki, Mark Lemley, Paul Kavanagh, Joanna X. Liang, Jianhang Zhou, Valerie Builoff, Jon Hainer, Sylvain Carre, Leanne Barrett, Andrew J. Einstein, Stacey Knight, Steve Mason, Viet T. Le, Wanda Acampa, Samuel Wopperer, Panithaya Chareonthaitawee, Daniel S. Berman, Marcelo F. Di Carli, Piotr J. Slomka

## Abstract

**Background:** Positron emission tomography (PET)/CT for myocardial perfusion imaging (MPI) provides multiple imaging biomarkers, often evaluated separately. We developed an artificial intelligence (AI) model integrating key clinical PET MPI parameters to improve the diagnosis of obstructive coronary artery disease (CAD).

**Methods:** From 17,348 patients undergoing cardiac PET/CT across four sites, we retrospectively enrolled 1,664 subjects who had invasive coronary angiography within 180 days and no prior CAD. Deep learning was used to derive coronary artery calcium score (CAC) from CT attenuation correction maps. XGBoost machine learning model was developed using data from one site to detect CAD, defined as left main stenosis ≥50% or ≥70% in other arteries. The model utilized 10 image-derived parameters from clinical practice: CAC, stress/rest left ventricle ejection fraction, stress myocardial blood flow (MBF), myocardial flow reserve (MFR), ischemic and stress total perfusion deficit (TPD), transient ischemic dilation ratio, rate pressure product, and sex. Generalizability was evaluated in the remaining three sites—chosen to maximize testing power and capture inter-site variability—and model performance was compared with quantitative analyses using the area under the receiver operating characteristic curve (AUC). Patient-specific predictions were explained using shapley additive explanations.

**Results:** There was a 61% and 53% CAD prevalence in the training (n=386) and external testing (n=1,278) set, respectively. In the external evaluation, the AI model achieved a higher AUC (0.83 [95% confidence interval (CI): 0.81-0.85]) compared to clinical score by experienced physicians (0.80 [0.77-0.82], p=0.02), ischemic TPD (0.79 [0.77–0.82], p<0.001), MFR (0.75 [0.72–0.78], p<0.001), and CAC (0.69 [0.66–0.72], p<0.001). The models’ performances were consistent in sex, body mass index, and age groups. The top features driving the prediction were stress/ischemic TPD, CAC, and MFR.

**Conclusion:** AI integrating perfusion, flow, and CAC scoring improves PET MPI diagnostic accuracy, offering automated and interpretable predictions for CAD diagnosis.

## Introduction

Positron emission tomography (PET) myocardial perfusion imaging (MPI), especially when combined with computed tomography (CT), plays a pivotal role in assessing patients with suspected coronary artery disease (CAD). The wealth of data provided by cardiac PET/CT includes (1) perfusion imaging, which depicts the extent of ischemia; (2) measurements of absolute myocardial blood flow (MBF), quantifying the volume of blood flow per unit time per myocardial mass (typically mL/min/g) and enhancing diagnostic accuracy beyond perfusion imaging alone^1–5^; (3) functional information; and (4) coronary artery calcium (CAC), a crucial indicator of atherosclerotic burden, all of which offer significant insights for risk stratification^6^. The latter can be derived automatically from CT attenuation correction (CTAC) scans which are routinely acquired as part of MPI exams on modern PET/CT systems^4, 7^.

However, currently, the assessment of CAD with PET/CT MPI does not optimally leverage the combined diagnostic power of these various imaging markers, including CAC^2, 3, 8, 9^. Recognizing this gap, we developed an artificial intelligence (AI) model that integrates a parsimonious set of 10 common PET MPI parameters—features often used intuitively but not systematically combined in clinical practice. While clinicians assess these parameters during interpretation, synthesizing all relevant data into a single interpretation can be challenging. In fact, there remains debate regarding the best methods to integrate just three features (CAC, relative perfusion, and blood flow)^10^, highlighting the need for an objective, automated approach.

Our AI approach harnesses deep learning and machine learning to comprehensively integrate and analyze key PET-CT parameters—including calcium burden, perfusion, myocardial blood flow (MBF), and functional metrics—enabling a more robust assessment of CAD. We based our model on standard PET imaging measurements rather than raw images to ensure generalizability, interpretability, and transparency—critical factors for clinical adoption. Furthermore, we rigorously validated our model on a large external testing cohort from three sites to confirm its robustness. To our knowledge, this work represents the first multicenter, externally validated, AI-driven cardiac PET MPI analysis.

## Methods

### Study population

In our multi-center study involving 17,348 patients undergoing cardiac PET/CT from the REgistry of Fast Myocardial Perfusion Imaging with NExt generation PET (REFINE PET), we retrospectively enrolled 1,664 patients from four sites who had suspected CAD, underwent ^82^Rubidium or ^13^N-ammonia PET MPI, and invasive coronary angiography within 180 days from the PET/CT (Supplementary Figure 1). CAD was defined as left main stenosis ≥50% or ≥70% in other epicardial arteries based on coronary catheterization and a review of the images by an experienced physician. Patients with prior myocardial infarction, percutaneous coronary intervention (PCI), and coronary artery bypass graft (CABG) were excluded from the analysis (Table 1). For patients who underwent several exams within the study period, only the initial exam was considered. Data from a single site, comprising 386 patients, were used for model training and optimization, while data from three additional sites, totaling 1,278 patients, were reserved for external testing. Clinical and demographic data were collected on the day of the MPI scan. Institutional review boards (IRB) approval was obtained at each site, and the study complies with the Declaration of Helsinki. Sites either obtained written informed consent or waiver of consent for the use of the de-identified data. To the extent allowed by data sharing agreements and IRB protocols, the data and code from this article will be shared upon written request.

**Table 1.** Patient characteristics.

|  | All patients,<br>n (%) | Training<br>dataset,<br>n (%) | External testing<br>dataset,<br>n (%) | P-value |
| --- | --- | --- | --- | --- |
| N (%) | 1,664 | 386 (23.2%) | 1,278 (76.8%) |  |
| Age, years | 68 (61, 75) | 71 (65, 80) | 67 (60, 74) | <0.001 |
| Male | 1,085 (65.2%) | 250 (64.8%) | 835 (65.3%) | 0.9 |
| Race |  |  |  | <0.001 |
| American Indian or Alaska Native | 11 (0.7%) | 1 (0.3%) | 10 (0.8%) |  |
| Asian | 36 (2.2%) | 16 (4.1%) | 20 (1.6%) |  |
| Black or African American | 102 (6.1%) | 59 (15.3%) | 43 (3.3%) |  |
| Native Hawaiian or Other Pacific<br>Islander | 22 (1.3%) | 2 (0.5%) | 20 (1.6%) |  |
| White | 1,410 (84.7%) | 297 (76.9%) | 1,113 (87.1%) |  |
| Unavailable | 83 (5.0%) | 11 (2.8%) | 72 (5.6%) |  |
| Smoking | 312 (18.8%) | 38 (9.8%) | 274 (21.4%) | <0.001 |
| BMI, kg/m<sup>2</sup> | 30 (26, 36) | 28 (24, 32) | 31 (26, 37) | <0.001 |
| Past medical history and comorbidities |  |  |  |  |
| Hypertension | 1,362 (81.9%) | 298 (77.2%) | 1,064 (83.3%) | 0.002 |
| Diabetes mellitus | 671 (40.3%) | 144 (37.3%) | 527 (41.2%) | 0.2 |
| Dyslipidemia | 1,198 (72.0%) | 243 (63.0%) | 955 (74.7%) | <0.001 |
| Family history of CAD | 561 (33.7%) | 58 (15.0%) | 503 (39.4%) | <0.001 |
| Peripheral Vascular Disease | 321 (19.3%) | 39 (10.1%) | 282 (22.1%) | <0.001 |
| Past Myocardial Infarction | 0 (0%) | 0 (0%) | 0 (0%) | NA |
| Past PCI | 0 (0%) | 0 (0%) | 0 (0%) | NA |
| CABG | 0 (0%) | 0 (0%) | 0 (0%) | NA |
| PET-CT acquisition parameters |  |  |  |  |
| Pharmacological Stress Agent |  |  |  | <0.001 |
| Dipyridamole | 19 (1.1%) | 1 (0.3%) | 18 (1.4%) |  |
| Adenosine | 131 (7.9%) | 42 (10.9%) | 89 (7.0%) |  |
| Regadenoson | 1,489 (89.5%) | 343 (88.9%) | 1,146 (89.7%) |  |
| Dobutamine | 25 (1.5%) | 0 (0.0%) | 25 (2.0%) |  |
| Isotope |  |  |  | <0.001 |
| <sup>13</sup>N-ammonia | 535 (32.2%) | 0 (0.0%) | 535 (41.9%) |  |
| <sup>82</sup>Rubidium | 1,129 (67.8%) | 386 (100.0%) | 743 (58.1%) |  |
| PET-CT quantitative image analysis parameters |  |  |  |  |
| Stress MBF, mL/g/min | 1.16 (0.77, 1.60) | 1.27 (0.86, 1.79) | 1.12 (0.75, 1.56) | <0.001 |
| MFR | 1.44 (1.03, 1.94) | 1.30 (0.89, 1.85) | 1.49 (1.08, 1.97) | <0.001 |
| Stress TPD | 9.9 (4.9, 18.3) | 8.9 (4.2, 16.8) | 10.4 (5.0, 18.9) | 0.012 |
| Stress TPD<5% | 434 (26.1%) | 118 (30.6%) | 316 (24.7%) | 0.026 |
| Ischemic TPD | 6.5 (3.4, 11.8) | 6.9 (3.3, 12.6) | 6.3 (3.4, 11.4) | 0.3 |
| TID | 1.05 (0.98, 1.13) | 1.05 (0.99, 1.16) | 1.05 (0.98, 1.12) | 0.095 |
| Stress EF, % | 62 (47, 72) | 63 (47, 72) | 62 (47, 72) | 0.6 |
| Rest EF, % | 61 (47, 70) | 62 (47, 70) | 61 (47, 70) | 0.6 |
| Coronary Artery Calcium Score |  |  |  | <0.001 |
| =0 | 346 (20.8%) | 58 (15.0%) | 288 (22.5%) |  |
| >0-100 | 289 (17.4%) | 58 (15.0%) | 231 (18.1%) |  |
| >100-400 | 343 (20.6%) | 79 (20.5%) | 264 (20.7%) |  |
| >400 | 686 (41.2%) | 191 (49.5%) | 495 (38.7%) |  |
| **Outcome** |  |  |  |  |
| Presence of obstructive CAD | 910 (54.7%) | 236 (61.1%) | 674 (52.7%) | 0.004 |

Categorical values were expressed as n (%). Continuous values were expressed as mean ± standard deviation or median (interquartile range), based on the Shapiro-Wilk test for normality. Variables were compared using a Pearson $\chi^2$ statistic for categorical variables and Mann-Whitney-Wilcoxon test for continuous variables.
Abbreviations: BMI – Body Mass Index; CABG – Coronary Artery Bypass Graft; CAD– Coronary Artery Disease; EF – Ejection Fraction; MBF – Myocardial Blood Flow; MFR – Myocardial Flow reserve; NA– Not Applicable; PCI– Percutaneous Coronary Intervention; TPD – Total Perfusion Deficit; TID – Transient Ischemic Dilation.

### PET Protocol

For all patients, same-day rest and pharmacologic stress ^82^Rubidium or ^13^N-ammonia PET MPI studies were conducted using a Biograph mCT 64 PET/CT scanner (Siemens Healthineers), a Discovery RX scanner (GE Healthcare), or a Discovery 710 scanner (GE Healthcare). A 6-minute rest list-mode acquisition was started immediately before administering weight-based doses of ^82^Rubidium or ^13^N-ammonia. Pharmacologic stress was induced using regadenoson (n=1,489), adenosine (n=131), dobutamine (n=25), or dipyridamole (n=19). Concurrent with the beginning of the injection, a 6-minute stress imaging acquisition was initiated. Prior to each PET acquisition for rest and stress, a low-dose helical CT scan was performed for attenuation correction, as detailed previously^11^.

### PET imaging and quantification

All PET imaging variables, including myocardial perfusion, blood flow, ejection fraction, and transient ischemic dilation (TID) ratio, were computed automatically in batch mode at the core laboratory with the dedicated software (QPET, Cedars-Sinai Medical Center, Los Angeles, CA)^12–14^. Rest and stress relative perfusion were quantified using total perfusion deficit (TPD). Normal myocardial perfusion was defined as stress TPD <5%^15^. Rest and stress MBF were measured using a 1-tissue compartment kinetic model for ^82^Rubidium PET and a 2-compartment model for ^13^N-ammonia PET^12–14^. MBF and the spillover fraction from the blood to the myocardium were determined via numeric optimization. Stress and rest flow values, expressed in units of mL/g/min, were computed for each sample on the polar map. Minimal segmental stress MBF (Stress MBF) was used in the AI model^16^. The rate–pressure product (RPP) was derived by multiplying the heart rate (bpm) by the systolic blood pressure (mmHg), which was then applied to normalize the rest MBF within the angiographic group using the formula (rest MBF × average RPP)/RPP. The average RPP value in the angiographic group was 8,500 bpm × mmHg^17^. Myocardial flow reserve (MFR) was computed as the ratio of minimal segmental stress MBF to minimal segmental rest MBF adjusted by RPP.

### Clinical scoring

PET/CT scans were visually assessed during clinical reporting by experienced physicians at each site, with knowledge of all available data, including stress and rest perfusion imaging, gated functional data, myocardial flow reserve, all other quantitative information, CT images and clinical information. The final visual assessment was performed by summed stress scores (SSS), summed rest scores (SRS), and summed difference score (SDS) using the 17-segment American Heart Association model. The SDS was used as the final clinical score, due to its reliability and standardization in summarizing myocardial perfusion abnormalities.

### AI Coronary Artery Calcium Scoring

We used our previously validated deep learning (DL) model for CAC segmentation and scoring^4, 18^. This model was trained and internally validated on data from three centers, including a total of 9,543 scans: 1,827 electrocardiographically gated CAC scans and 7,716 CTAC maps^4^. Using the established deep learning segmentation method, we automatically derived CAC scores from CTAC maps.

### Classification Model and internal model testing

Extreme Gradient Boosting (XGBoost) models (version 1.7.3), a leading machine learning approach, were used for CAD diagnosis^19^. Initially, 10-fold cross-validation was applied across the training dataset, where each fold allocated 90% of the data for model training and the remaining 10% for internal validation. To optimize model performance, hyper-parameter tuning was conducted within each fold through grid search, selecting the optimal hyper-parameter configuration from all tested combinations. In each 10-fold subset, an internal 10-fold cross-validation regimen further allocated 90% of the subset data for fitting and 10% for tuning, maximizing training data utilization and mitigating overfitting (Supplementary Figure 2).

For the subsequent model evaluation phase, an external testing set was used. This additional testing set, originating from three distinct sites from that of the training data, was employed to rigorously assess the predictive performance on unseen data, providing an unbiased evaluation. The final model tested was created using the optimal hyper-parameters obtained through grid search, retrained on the entire training set, and then tested on this external testing set.

The diagnostic performance of the holistic AI model was evaluated using receiver operating characteristic area under the curve (AUC) and sensitivity analysis. Sensitivity was compared by adjusting prediction thresholds to match specificity across methods, then assessing the corresponding sensitivity. Comparisons focused on high-risk groups in external testing set, defined by Duke category 6 (≥50% left main stenosis, ≥70% 3-vessel disease, or 2-vessel CAD involving proximal LAD) and Duke category 5 (2-vessel severe stenosis excluding proximal LAD, one-vessel severe stenosis in proximal LAD, or ≥50% 3-vessel moderate stenosis)^20^. Quantitative analysis include minimum 3-vessel MFR <2^21^, minimum 3-vessel MBF <1.8^22^, and ischemic TPD >5%^23^.

Additionally, subgroup analyses were conducted to gain further insights into the performance of the AI models. Subgroups were categorized based on sex (male, female), age (<65, ≥65 years), and body mass index (BMI<30 kg/m^2^, BMI≥30kg/m^2^)^24^. Due to limited data for minority races, the race-based subgroup analysis was excluded.

## Models

Six models were used for the CAD diagnosis: 1 – CAC derived from CTAC maps, 2 – ischemic TPD, 3 – Stress MBF, 4 – MFR, 5 – Summed Difference Score (clinical score) obtained during clinical reading by an experienced physician, 6 – AI, which employs solely PET/CT derived data without incorporating additional clinical variables. The AI model incorporated the CAC along with: Stress MBF, MFR, ischemic TPD, stress TPD, stress and rest left ventricular ejection fraction (LVEF), RPP, TID, and sex, resulting in a total of ten image-derived features to mimic clinical practice (Table 2). The model is designed to hold regardless of age, BMI, obesity, or other demographic factors.

**Table 2.**
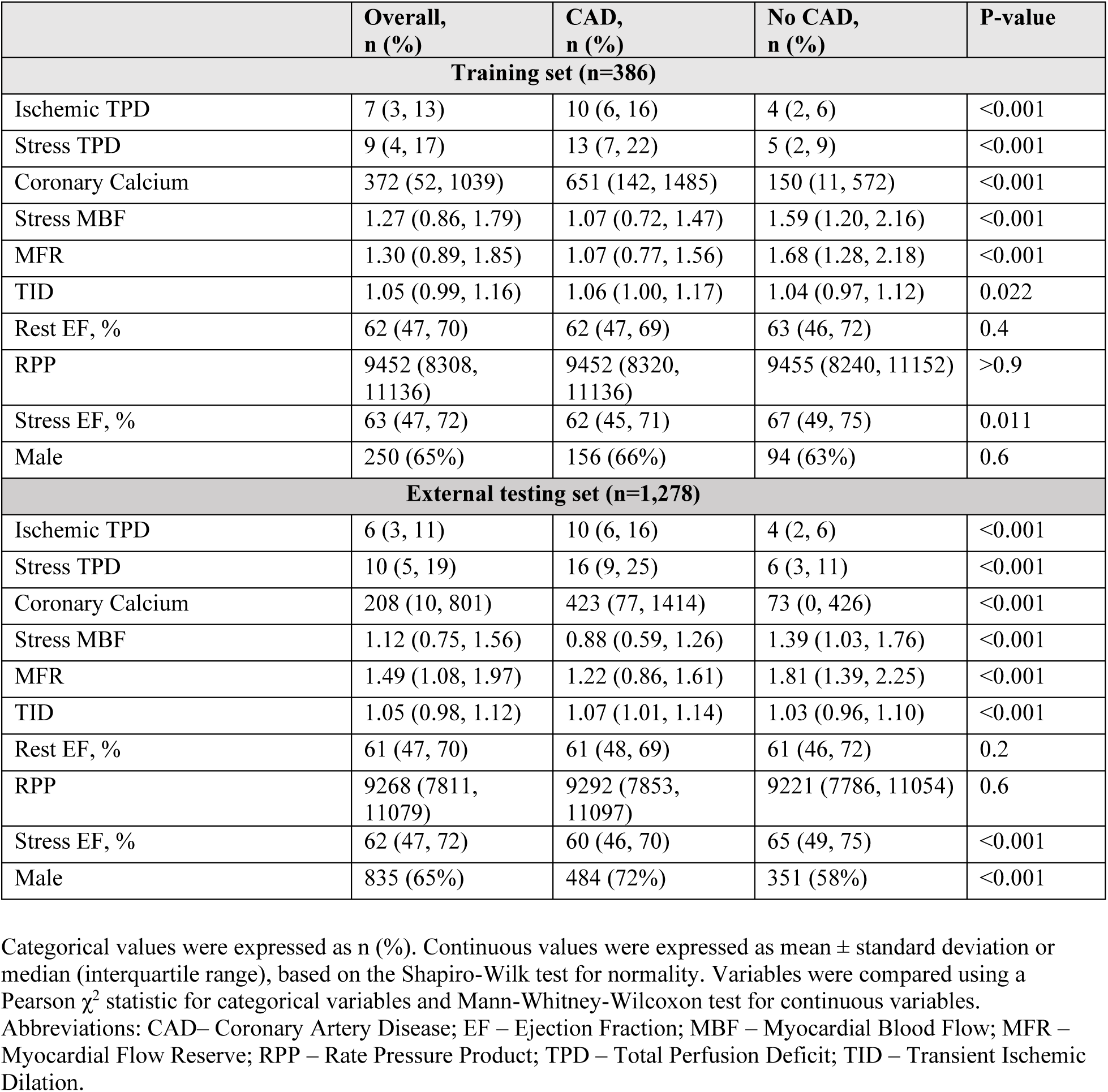
Parameters included in the AI model from the training set (n=386) and external testing set (n=1,278).

|  | Overall,<br>n (%) | CAD,<br>n (%) | No CAD,<br>n (%) | P-value |
| --- | --- | --- | --- | --- |
| **Training set (n=386)** |  |  |  |  |
| Ischemic TPD | 7 (3, 13) | 10 (6, 16) | 4 (2, 6) | <0.001 |
| Stress TPD | 9 (4, 17) | 13 (7, 22) | 5 (2, 9) | <0.001 |
| Coronary Calcium | 372 (52, 1039) | 651 (142, 1485) | 150 (11, 572) | <0.001 |
| Stress MBF | 1.27 (0.86, 1.79) | 1.07 (0.72, 1.47) | 1.59 (1.20, 2.16) | <0.001 |
| MFR | 1.30 (0.89, 1.85) | 1.07 (0.77, 1.56) | 1.68 (1.28, 2.18) | <0.001 |
| TID | 1.05 (0.99, 1.16) | 1.06 (1.00, 1.17) | 1.04 (0.97, 1.12) | 0.022 |
| Rest EF, % | 62 (47, 70) | 62 (47, 69) | 63 (46, 72) | 0.4 |
| RPP | 9452 (8308, 11136) | 9452 (8320, 11136) | 9455 (8240, 11152) | >0.9 |
| Stress EF, % | 63 (47, 72) | 62 (45, 71) | 67 (49, 75) | 0.011 |
| Male | 250 (65%) | 156 (66%) | 94 (63%) | 0.6 |
| **External testing set (n=1,278)** |  |  |  |  |
| Ischemic TPD | 6 (3, 11) | 10 (6, 16) | 4 (2, 6) | <0.001 |
| Stress TPD | 10 (5, 19) | 16 (9, 25) | 6 (3, 11) | <0.001 |
| Coronary Calcium | 208 (10, 801) | 423 (77, 1414) | 73 (0, 426) | <0.001 |
| Stress MBF | 1.12 (0.75, 1.56) | 0.88 (0.59, 1.26) | 1.39 (1.03, 1.76) | <0.001 |
| MFR | 1.49 (1.08, 1.97) | 1.22 (0.86, 1.61) | 1.81 (1.39, 2.25) | <0.001 |
| TID | 1.05 (0.98, 1.12) | 1.07 (1.01, 1.14) | 1.03 (0.96, 1.10) | <0.001 |
| Rest EF, % | 61 (47, 70) | 61 (48, 69) | 61 (46, 72) | 0.2 |
| RPP | 9268 (7811, 11079) | 9292 (7853, 11097) | 9221 (7786, 11054) | 0.6 |
| Stress EF, % | 62 (47, 72) | 60 (46, 70) | 65 (49, 75) | <0.001 |
| Male | 835 (65%) | 484 (72%) | 351 (58%) | <0.001 |

Categorical values were expressed as n (%). Continuous values were expressed as mean ± standard deviation or median (interquartile range), based on the Shapiro-Wilk test for normality. Variables were compared using a Pearson $\chi^2$ statistic for categorical variables and Mann-Whitney-Wilcoxon test for continuous variables. Abbreviations: CAD– Coronary Artery Disease; EF – Ejection Fraction; MBF – Myocardial Blood Flow; MFR – Myocardial Flow Reserve; RPP – Rate Pressure Product; TPD – Total Perfusion Deficit; TID – Transient Ischemic Dilation.

### Model Explainability

The predictive power of model variables was assessed using XGBoost feature importance, measured by information gain to quantify accuracy improvement from each feature^19^. Features were ranked by overall importance, with the most influential at the top. SHapley Additive exPlanations (SHAP), a game-theoretic method, explained how features contributed to individual predictions^25^. In SHAP plots, dots represent the SHAP value of each patient for a feature, with positive values increasing predictions and negative values decreasing them. The x-axis distance from zero indicates the magnitude of influence.

### Statistical Analysis

We assessed the distribution of data using the Shapiro-Wilk test. Categorical variables were reported as n (%) and continuous variables with a normal distribution as mean ± standard deviation (SD), while non-normal distribution was reported as median with interquartile range (IQR) [IQ1-IQ3]. We used the Pearson’s χ2 test to assess the differences between categorical variables, the student’s t-test for continuous variables with normal distribution, the Mann-Whitney-Wilcoxon test for non-parametric continuous variables. Predictions of CAD by ischemia, stress MBF, MFR, coronary calcium, and AI models were assessed by pairwise comparisons of the AUC with the DeLong test^26^. A two-tailed p-value of <0.05 was considered statistically significant. All statistical analyses were performed with Pandas (version 2.1.1), Numpy (version 1.24.3), Scipy (version 1.11.4), Lifelines (version 0.28.0) and Scikit-learn (version 1.3.0) in Python 3.11.5 (Python Software Foundation, Wilmington, DE, USA), as well as “nricens” package (version 1.6) in R version 4.3.2 (R Foundation for Statistical Computing, Vienna, Austria).

## Results

### Study population

Patient characteristics are shown in Table 1. Among the 1,664 participants, 1,085 (65%) were male, with a median age of 68 years (IQR 61, 75). Of these patients, 386 were allocated for model training and optimization, while 1,278 from separate institutions were used for external testing. The training data were complete, containing no missing values. The prevalence of CAD was significantly higher in the internal training set than in the external testing set (61% vs. 53%, p=0.004).

### Myocardial imaging analysis parameters

In both the internal training and external testing sets, patients with CAD demonstrated significantly higher median ischemic TPD, stress TPD, and TID compared to those without CAD (p<0.001 for all in the testing set; p<0.001 for TPDs and p=0.022 for TID in the training set) (Table 2). Additionally, patients with CAD had notably lower stress MBF (p<0.001), MFR (p<0.001), and median stress ejection fraction (p=0.011 in training, p<0.001 in testing).

### Coronary Artery Calcium

Of the internal training cohort (n=386), CAC was 0 in 58 (15.0%) patients, >0-100 in 58 (15.0%), >100-400 in 79 (20.5%), and CAC >400 in 191 (49.5%) subjects. In the external testing set (n=1,278), 288 (22.5%) had a CAC of 0, 231 (18.1%) had a CAC between >0-100, 264 (20.7%) had a CAC between >100-400, and 495 (38.7%) had a CAC >400 (Table 1).

### Model Performance

In external testing, we compared the AI model with clinical measurements (Figure 1). The comprehensive machine learning model (AUC 0.83, [0.81-0.85]) outperformed clinical score from experienced physicians (0.80 [0.77-0.82], p=0.02), ischemic TPD (0.79, [0.77-0.82], p<0.001), MFR (0.75, [0.72-0.78], p<0.001), stress MBF (0.75, [0.72-0.77], p<0.001), and CAC (0.69, [0.66-0.72], p<0.001) (Figure 2 left panel, Supplementary Table 1). Figure 2 (right panel) shows that the AI model achieved significantly higher sensitivity (*p* < 0.001) than quantitative thresholds including minimum 3-vessel MFR <2, MBF <1.8, and ischemic TPD ≥5%, when matched for specificity in the external testing set. This improvement was driven by the model’s ability to identify more high-risk patients classified by Duke 6 and Duke 5 criteria, compared to quantitative measurements at matched specificity (Supplementary Table 2). These results suggest that a holistic AI-based approach not only improves overall diagnostic sensitivity but also enables more accurate identification of high-risk patients who may be missed by individual quantitative metrics.

**Figure 1.**
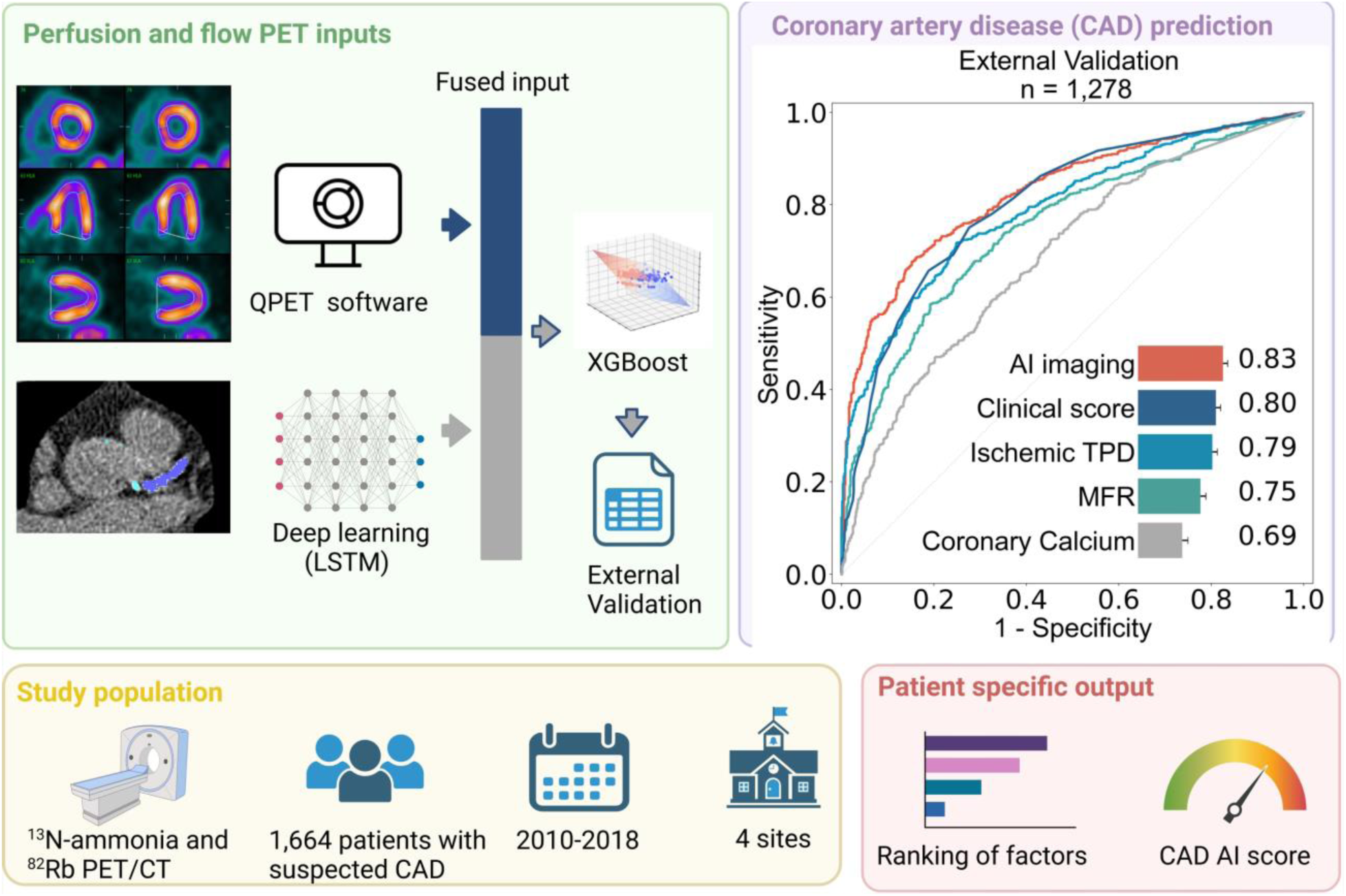
Central illustration. Artificial intelligence (AI) model integrating fully automated PET flow, perfusion quantification, and deep learning (DL)-based coronary artery calcium in all patients undergoing myocardial perfusion imaging positron emission computed tomography/computed tomography (PET/CT). Receiver-operating characteristics curve for coronary artery disease (CAD) diagnosis and area under the receiver-operating characteristic curve values of AI model vs ischemic total perfusion deficit (TPD), myocardial flow reserve (MFR), and coronary calcium in external validation assessment (n=1,278). Created in BioRender. Zhang, W. (2025) https://BioRender.com/y27z326. Abbreviations: LSTM – Long Short-Term Memory Network; XGBoost – Extreme Gradient Boosting.

**Figure 2.**
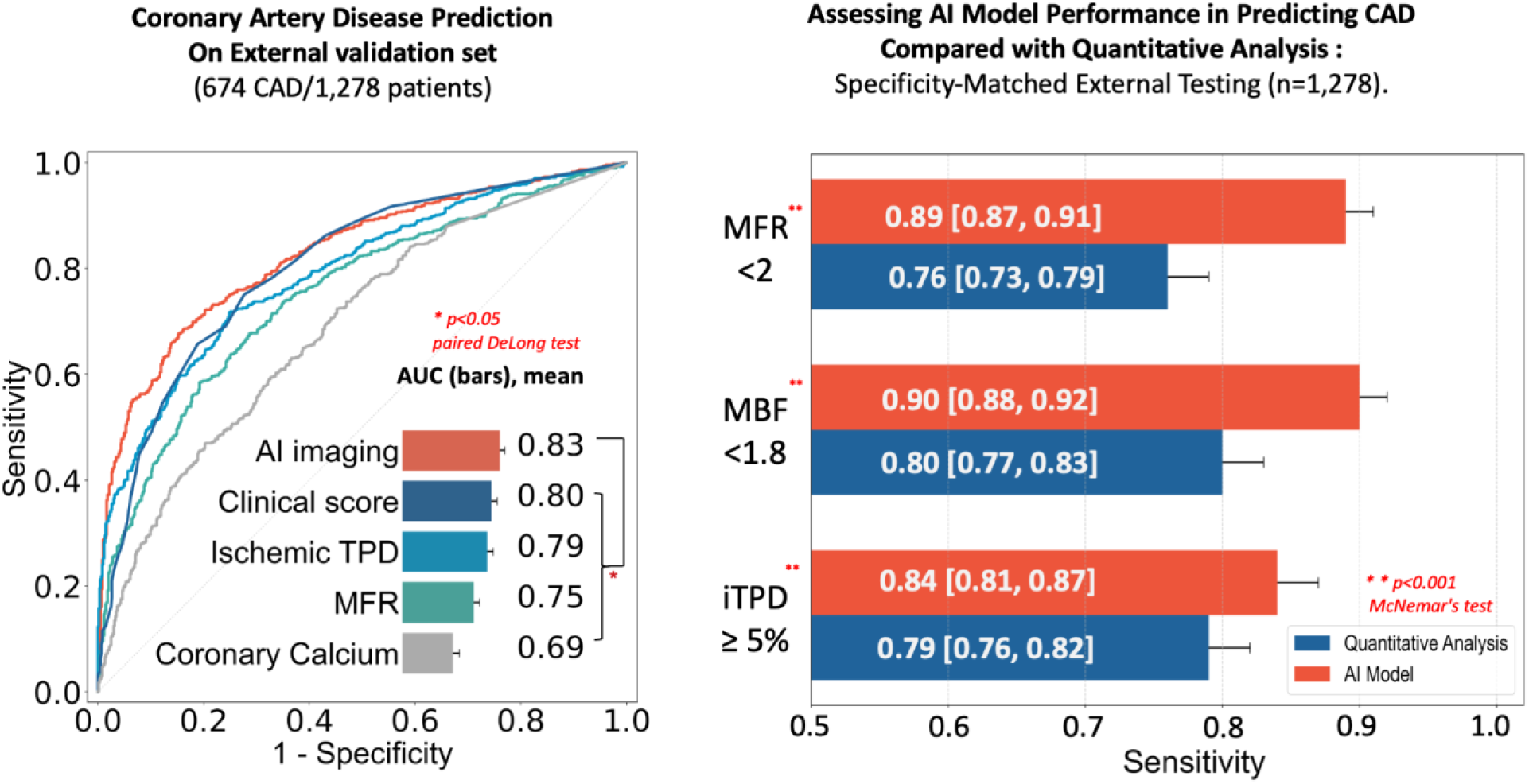
AI Model Performance and Feature Importance for CAD Prediction. **Left panel**: Receiver-operating characteristics curve for coronary artery disease (CAD) and areas under the curve (AUC) values. AI Imaging model incorporates sex, coronary artery calcium score, rest and stress left ventricle ejection fraction (LVEF), rate pressure product (RPP), myocardial blood flow (MBF) and flow reserve (MFR), and perfusion measurements derived from PET imaging. **Right panel**: Sensitivity analysis comparing AI and quantitative methods for predicting CAD. Performance assessed on the external validation set, with specificity matched between AI predictions and quantitative analysis. MFR and MBF were selected from the minimum of the 3 vessels. Notably, AI model shows significant improvement over MFR, MBF, and iTPD in terms of sensitivity. Abbreviations: AI – Artificial Intelligence; TID – Transient Ischemic Dilation; iTPD – Ischemic Total Perfusion Deficit.

The top features driving the prediction were ischemic and stress TPD, CAC, and MFR using SHAP values (Supplementary Figure 3). In a predefined subgroup analysis, the AI models exhibited comparable performance across various subgroups: female and male patients (AUC 0.83 vs 0.83, p=0.65), patients classified as non-obese (BMI < 30kg/m^2^) and obese (BMI ≥30kg/m^2^) (AUC 0.84 vs 0.81, p=0.10), and older (age ≥ 65 years) and younger (age < 65 years) (AUC 0.84 vs 0.81, p=0.36) (Supplementary Figure 4).

### Individualized Inference by the AI Model

In Figure 3, two cases illustrate the ability of the AI model to integrate multiple PET MPI parameters for CAD diagnosis. Case A demonstrates how the model accurately identifies of obstructive CAD, emphasizing MFR as a key driver. Case B highlights the capability of the model to correctly rule out CAD despite borderline TPD and clinician-assigned stress scores. These examples demonstrate the model’s effectiveness in leveraging integrated biomarkers to align predictions with patient outcomes.

**Figure 3.**
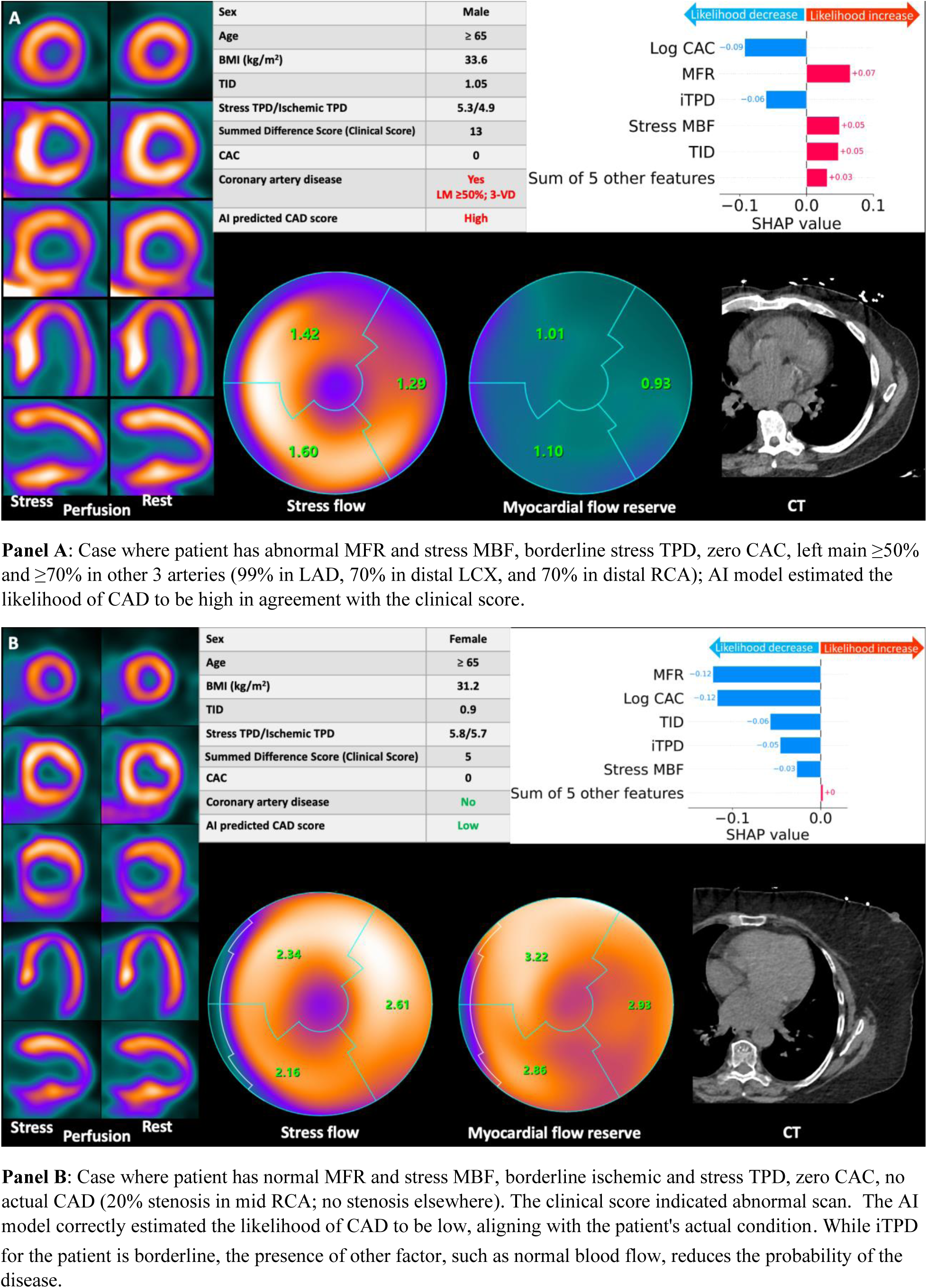
Examples of PET/CT Imaging in Predicted CAD Patients. Examples of patients undergoing PET/CT myocardial perfusion imaging predicted to have CAD. Bar plots demonstrate the top 5 parameters with the highest influence on per-patient prediction of CAD. Abbreviations: BMI – Body Mass Index; CAC – Coronary Calcium Score; CAD – Coronary Artery Disease; CT – Computed Tomography; iTPD – Ischemic TPD; LCX – Left Circumflex Artery; LAD – Left Anterior Descending; LM – Left Main; LVEF – Left Ventricular Ejection Fraction; MBF – Myocardial Blood Flow; MFR – Myocardial Flow Reserve; PET – Positron Emission Tomography; RPP – Rate Pressure Product; TID – Transient Ischemic Dilation; TPD – Total Perfusion Deficit; VD – Vessel Disease.

## Discussion

PET/CT, with its unique ability to quantify absolute MBF, allows robust detection of obstructive coronary stenoses across the spectrum of CAD including diffuse atherosclerosis, microvascular disease or multivessel disease. Consequently, PET is the fastest growing non-invasive cardiac imaging test^27^. However, to take full advantage of PET/CT it is necessary to combine complex data, which is currently not performed systematically.

In this study, we developed, tested, and externally validated an interpretable AI model that integrates key quantitative PET MPI parameters—including stress/rest perfusion, myocardial flow, ejection fraction, and AI-derived calcium scores, sex—into a unified assessment. In multicenter external validation, our AI approach outperformed both expert physicians (provided with complete imaging/clinical data during assessment) and conventional quantitative methods. The AI model demonstrated improved sensitivity and superior detection of high-risk patients highlighting its strong generalizability despite inter-center variability. To avoid the perception of AI as a ‘black box’ we have provided both patient-level and model-level explanation by highlighting the most influential factors in predicting significant CAD, enhancing physician understanding and confidence in AI-driven predictions. Collectively, these characteristics underscore the tremendous potential of our AI model for real-world practical approach to diagnosis incorporating all major factors currently considered by physicians, including calcium derived by deep learning from CT attenuation maps.

To date, several single site studies have investigated benefits integrating subsets of PET/CT MPI parameters,^2, 3, 9, 21, 23, 28–30^. Initial studies focused on optimizing risk stratification by combining MBF and perfusion data^2, 21, 30^. Gupta et al. established that MFR serves as a stronger predictor of cardiac mortality than maximal MBF in patients with stable CAD, with integrated assessment of MFR and MBF identifying distinct prognostic risk profiles, particularly highlighting MFR impairment as a key mortality indicator^9^. Gould et al. proposed the coronary flow capacity measure, which integrates regional stress MBF and MFR into one variable^28^. Using data from a single center, Singh et al. developed a explainable deep learning model for all-cause mortality prediction, integrating polar maps of stress and rest perfusion, MBF, MFR, and spill-over fraction combined with cardiac volumes, singular indices, and sex^29^. These prior studies have consistently demonstrated that combing multiparametric PET data improves risk stratification with more comprehensive models outperforming approaches relying on only two or three variables.

Few studies have attempted to improve diagnostic accuracy of cardiac PET/CT through such an integrative approach^2, 6, 29^. Poitrasson-Rivière et al. demonstrated that combining MFR and relative perfusion improves the detection of obstructive CAD^3^. Beyond flow and perfusion measures, assessments of the coronary atherosclerotic burden can provide additional information. Zampella et al. demonstrated that combining CAC, MBF and MFR provides incremental information about the presence of CAD^31^. Similarly, Brodov et al. showed that integrating per-vessel ischemic TPD with CAC improves CAD detection^23^. While previous studies combining PET and CAC data relied on dedicated ECG-gated CT and manual expert analysis, in this study we obtained CAC data automatically from PET CTAC. By leveraging CTAC for disease burden assessments our approach can be employed for all PET MPI studies rather than being limited to exams which include a dedicated CAC scan. Our study is the first AI empowered multicenter PET analysis with rigorous external validation. While previous efforts relied on single center data and hence employed prospective validation, we developed our model using data from one site and tested its performance on unseen data from three independent high-volume PET centers. This provides critical evaluation of the external generalizability of our approach which is further strengthened by the utilization of PET imaging measurements which vary less between institution compared to actual PET images^32^.

Traditional clinical studies combining multiple PET parameters required separate threshold values for key parameters and relied on conventional statistics ^2, 21^. In contrast, our AI model integrates multidimensional data as continuous variables, delivering a probability-based assessment of obstructive CAD without relying on arbitrary thresholds. Using SHAP values, the model highlights parameters influencing CAD predictions for individual patients (Figure 3), enabling physicians to validate and understand case-specific diagnostic factors. This explainable AI model enhances clinical workflows by providing quantitative, automated, and objective insights, ultimately improving the efficiency of PET/CT MPI-based CAD prediction. By offering patient-specific diagnostic insights, it empowers physicians with comprehensive and actionable decision support at the point of care.

### Limitations

This study has a few limitations. While it was a retrospective study that only included four sites, it represents the largest PET/MPI and CAC study with invasive angiography data used as a gold standard. Nevertheless, further external testing of the developed model should be conducted in the future. While most processing was automated there was a quality control step of myocardial contours performed by an experienced technologist. Additionally, although invasive angiography serves as a traditional diagnostic endpoint, it is important to acknowledge the possibility of significant disease being present despite negative angiographic findings. For example, flow assessment by PET MPI can detect microvascular dysfunction and diffuse atherosclerosis without significant angiographic stenosis, an aspect that was not addressed in the current study. Finally, race analysis was excluded from this study due to the limited availability of data on individuals from minority racial groups, such as Black individuals.

### Conclusions

This study introduces a novel AI model that integrates PET MPI parameters with deep learning-derived calcium scores, significantly enhancing CAD diagnosis. It outperforms experienced physicians, offers robust and interpretable assessments, and represents the first multicenter study with external validation for AI-driven cardiac PET MPI analysis.

## List of abbreviation

AI: artificial intelligence
AUC: area under the receiver-operating characteristic curve
BMI: body mass index
CABG: coronary artery bypass graft
CAC: coronary artery calcium
CAD: coronary artery disease
CT: computed tomography
CTAC: computed tomography attenuation correction
DL: deep learning
LVEF: left ventricle ejection fraction
MBF: myocardial blood flow
MFR: myocardial flow reserve
MPI: myocardial perfusion imaging
PCI: percutaneous coronary intervention
PET: positron emission computed tomography
SHAP: shapley additive explanations
TID: transient ischemic dilation ratio
TPD: total perfusion deficit

## Contributors

WZ conducted data processing/experiments/analysis, and co-wrote the manuscript. JK co-wrote the manuscript and contributed to materials, clinical expertise. PJS designed the study, provided the overall guidance and study funding, co-wrote the manuscript, and contributed materials, clinical expertise, and technical expertise. AS, GR, JY, DH, RJM, DD, GD, GK, ML, PBK, JXL, JZ, VB, JH, SC, LB, AJE, SK, SM, VTL, WA, SW, PC, DSB, and MFDC contributed to materials, clinical expertise, and technical expertise. All authors critically revised the manuscript and contributed to its formation. All authors had full access to all the data in the study, accept the final responsibility to submit for publication and take responsibility for the contents of the manuscript.

## Competing interests

Dr. Robert Miller received consulting fees and research support from Pfizer. Drs. Berman and Slomka, and Paul B. Kavanagh participate in software royalties for QPS software at Cedars-Sinai Medical Center. Dr. Slomka has received consulting fees from Synektik. Dr. Berman has served as a consultant for GE Healthcare. Dr. Einstein reports receiving a speaker’s fee from Ionetix, consulting fees from W. L. Gore & Associates and Artrya, authorship fees from Wolters Kluwer Healthcare—UpToDate, and serving on a scientific advisory board for Canon Medical Systems USA; his institution has grants/grants pending from Attralus, BridgeBio, Canon Medical Systems USA, GE HealthCare, Intellia Therapeutics, Ionis Pharmaceuticals, Mediwhale, Neovasc, Pfizer, Roche Medical Systems, and W. L. Gore & Associates. Dr. Di Carli reports on institutional research grants from Gilead Sciences, Xylocor, Sun Pharma, Intellia Therapeutics, Alnylam Pharmaceuticals, and Amgen. He also receives consulting fees from MedTrace, Valo Health, and IBA. The remining authors have nothing to disclose.

## Data and code sharing

To the extent allowed by data and code sharing agreements and IRB protocols, the deidentified data and analysis code from this manuscript will be shared upon written request. The analysis code will be made available on GitHub: https://github.com/qimagingAI/AI4CADdiagnosisPET-CT.

## Acknowledgements

This research was supported in part by grant R35HL161195 from the National Heart, Lung, and Blood Institute of the National Institutes of Health and grant R01EB034586 from the National Institute of Biomedical Imaging and Bioengineering of the National Institutes of Health. The content is solely the responsibility of the authors and does not necessarily represent the official views of the National Institutes of Health.

## Notes

### Author Declarations

Institutional review boards (IRB) approval was obtained at each site, and the study complies with the Declaration of Helsinki. Sites either obtained written informed consent or waiver of consent for the use of the de-identified data.

## References

1. Di Carli MF, Dorbala S and Hachamovitch R. Integrated cardiac PET-CT for the diagnosis and management of CAD. J Nucl Cardiol 2006; 13: 139–144. DOI: 10.1007/BF02971234.

2. Murthy VL, Naya M, Foster CR, et al. Improved cardiac risk assessment with noninvasive measures of coronary flow reserve. Circulation 2011; 124: 2215–2224. 20111017. DOI: 10.1161/CIRCULATIONAHA.111.050427.

3. Poitrasson-Riviere A, Moody JB, Renaud JM, et al. Integrated myocardial flow reserve (iMFR) assessment: optimized PET blood flow quantification for diagnosis of coronary artery disease. Eur J Nucl Med Mol Imaging 2023; 51: 136–146. 20231009. DOI: 10.1007/s00259-023-06455-2.

4. Pieszko K, Shanbhag A, Killekar A, et al. Deep Learning of Coronary Calcium Scores From PET/CT Attenuation Maps Accurately Predicts Adverse Cardiovascular Events. JACC Cardiovasc Imaging 2023; 16: 675–687. 2022/10/27. DOI: 10.1016/j.jcmg.2022.06.006.

5. McCubrey RO, Mason SM, Le VT, et al. A highly predictive cardiac positron emission tomography (PET) risk score for 90-day and one-year major adverse cardiac events and revascularization. J Nucl Cardiol 2023; 30: 46–58. 20221219. DOI: 10.1007/s12350-022-03028-y.

6. van Velzen SGM, Dobrolinska MM, Knaapen P, et al. Automated cardiovascular risk categorization through AI-driven coronary calcium quantification in cardiac PET acquired attenuation correction CT. Journal of Nuclear Cardiology 2023; 30: 955–969. DOI: 10.1007/s12350-022-03047-9.

7. Anderson JL, Knight S, McCubrey RO, et al. Absent or Mild Coronary Calcium Predicts Low-Risk Stress Test Results and Outcomes in Patients Considered for Flecainide Therapy. J Cardiovasc Pharmacol Ther 2021; 26: 648–655. 20210921. DOI: 10.1177/10742484211046671.

8. Maddahi J, Agostini D, Bateman TM, et al. Flurpiridaz F-18 PET Myocardial Perfusion Imaging in Patients With Suspected Coronary Artery Disease. J Am Coll Cardiol 2023; 82: 1598–1610. DOI: 10.1016/j.jacc.2023.08.016.

9. Gupta A, Taqueti VR, van de Hoef TP, et al. Integrated Noninvasive Physiological Assessment of Coronary Circulatory Function and Impact on Cardiovascular Mortality in Patients With Stable Coronary Artery Disease. Circulation 2017; 136: 2325–2336. 20170901. DOI: 10.1161/CIRCULATIONAHA.117.029992.

10. Mannarino T, D’Antonio A, Assante R, et al. Combined evaluation of CAC score and myocardial perfusion imaging in patients at risk of cardiovascular disease: where are we and what do the data say. J Nucl Cardiol 2023; 30: 2349–2360. 20230510. DOI: 10.1007/s12350-023-03288-2.

11. Bacharach SL. PET/CT attenuation correction: breathing lessons. J Nucl Med 2007; 48: 677–679. DOI: 10.2967/jnumed.106.037499.

12. Dekemp RA, Declerck J, Klein R, et al. Multisoftware reproducibility study of stress and rest myocardial blood flow assessed with 3D dynamic PET/CT and a 1-tissue-compartment model of 82Rb kinetics. J Nucl Med 2013; 54: 571–577. 2013/03/01. DOI: 10.2967/jnumed.112.112219.

13. Nakazato R, Berman DS, Dey D, et al. Automated quantitative Rb-82 3D PET/CT myocardial perfusion imaging: normal limits and correlation with invasive coronary angiography. J Nucl Cardiol 2012; 19: 265–276. 20111228. DOI: 10.1007/s12350-011-9496-3.

14. Slomka PJ, Alexanderson E, Jácome R, et al. Comparison of clinical tools for measurements of regional stress and rest myocardial blood flow assessed with 13N-ammonia PET/CT. J Nucl Med 2012; 53: 171–181. 2012/01/10. DOI: 10.2967/jnumed.111.095398.

15. Otaki Y, Betancur J, Sharir T, et al. 5-Year Prognostic Value of Quantitative Versus Visual MPI in Subtle Perfusion Defects: Results From REFINE SPECT. JACC Cardiovasc Imaging 2020; 13: 774–785. 2019/06/17. DOI: 10.1016/j.jcmg.2019.02.028.

16. Otaki Y, Van Kriekinge SD, Wei C-C, et al. Improved myocardial blood flow estimation with residual activity correction and motion correction in 18F-flurpiridaz PET myocardial perfusion imaging. European Journal of Nuclear Medicine and Molecular Imaging 2022; 49: 1881–1893. DOI: 10.1007/s00259-021-05643-2.

17. Kuronuma K, Wei CC, Singh A, et al. Automated Motion Correction for Myocardial Blood Flow Measurements and Diagnostic Performance of (82)Rb PET Myocardial Perfusion Imaging. J Nucl Med 2024; 65: 139–146. 2023/12/05. DOI: 10.2967/jnumed.123.266208.

18. Williams MC, Shanbhag AD, Zhou J, et al. Automated vessel-specific coronary artery calcification quantification with deep learning in a large multi-centre registry. Eur Heart J Cardiovasc Imaging 2024; 25: 976–985. 2024/02/20. DOI: 10.1093/ehjci/jeae045.

19. Chen T and Guestrin C. XGBoost: A Scalable Tree Boosting System. Proceedings of the 22nd ACM SIGKDD International Conference on Knowledge Discovery and Data Mining. San Francisco, California, USA: Association for Computing Machinery, 2016, p. 785–794.

20. Reynolds HR, Shaw LJ, Min JK, et al. Outcomes in the ISCHEMIA Trial Based on Coronary Artery Disease and Ischemia Severity. Circulation 2021; 144: 1024–1038. 20210909. DOI: 10.1161/CIRCULATIONAHA.120.049755.

21. Ziadi MC, Dekemp RA, Williams KA, et al. Impaired myocardial flow reserve on rubidium-82 positron emission tomography imaging predicts adverse outcomes in patients assessed for myocardial ischemia. J Am Coll Cardiol 2011; 58: 740–748. DOI: 10.1016/j.jacc.2011.01.065.

22. Hajjiri MM, Leavitt MB, Zheng H, et al. Comparison of positron emission tomography measurement of adenosine-stimulated absolute myocardial blood flow versus relative myocardial tracer content for physiological assessment of coronary artery stenosis severity and location. JACC Cardiovasc Imaging 2009; 2: 751–758. DOI: 10.1016/j.jcmg.2009.04.004.

23. Brodov Y, Gransar H, Dey D, et al. Combined Quantitative Assessment of Myocardial Perfusion and Coronary Artery Calcium Score by Hybrid 82Rb PET/CT Improves Detection of Coronary Artery Disease. J Nucl Med 2015; 56: 1345–1350. 2015/07/15. DOI: 10.2967/jnumed.114.153429.

24. Purnell JQ. Definitions, Classification, and Epidemiology of Obesity. In: Feingold KR, Anawalt B, Blackman MR, et al. (eds) Endotext. South Dartmouth (MA), 2000.

25. Lundberg SM and Lee S-I. A unified approach to interpreting model predictions. Proceedings of the 31st International Conference on Neural Information Processing Systems. Long Beach, California, USA: Curran Associates Inc., 2017, p. 4768–4777.

26. Delong ER, Delong DM and Clarkepearson DI. Comparing the Areas under 2 or More Correlated Receiver Operating Characteristic Curves - a Nonparametric Approach. Biometrics 1988; 44: 837–845. DOI: Doi 10.2307/2531595.

27. Reeves RA, Halpern EJ and Rao VM. Cardiac Imaging Trends from 2010 to 2019 in the Medicare Population. Radiology: Cardiothoracic Imaging 2021; 3: e210156. DOI: 10.1148/ryct.2021210156.

28. Gould KL, Kitkungvan D, Johnson NP, et al. Mortality Prediction by Quantitative PET Perfusion Expressed as Coronary Flow Capacity With and Without Revascularization. JACC: Cardiovascular Imaging 2021; 14: 1020–1034. DOI: 10.1016/j.jcmg.2020.08.040.

29. Singh A, Kwiecinski J, Miller RJH, et al. Deep Learning for Explainable Estimation of Mortality Risk From Myocardial Positron Emission Tomography Images. Circ Cardiovasc Imaging 2022; 15: e014526. 20220920. DOI: 10.1161/CIRCIMAGING.122.014526.

30. Valenta I and Schindler TH. PET-determined myocardial perfusion and flow in coronary artery disease characterization. Journal of Medical Imaging and Radiation Sciences 2024; 55: S44–S50. DOI: 10.1016/j.jmir.2024.02.010.

31. Zampella E, Acampa W, Assante R, et al. Combined evaluation of regional coronary artery calcium and myocardial perfusion by (82)Rb PET/CT in predicting lesion-related outcome. Eur J Nucl Med Mol Imaging 2020; 47: 1698–1704. 20191214. DOI: 10.1007/s00259-019-04534-x.

32. Akamatsu G, Tsutsui Y, Daisaki H, et al. A review of harmonization strategies for quantitative PET. Ann Nucl Med 2023; 37: 71–88. 20230106. DOI: 10.1007/s12149-022-01820-x.

