## Supplemental Material for "Multicenter Evaluation of Interpretable AI for Coronary Artery Disease Diagnosis from PET Biomarkers"

### Supplemental Materials

| | |
|---|---|
| Supplementary Table 1 | Page 2 |
| Supplementary Table 2 | Page 3 |
| Supplementary Figure 1 | Page 4 |
| Supplementary Figure 2 | Page 5 |
| Supplementary Figure 3 | Page 6 |
| Supplementary Figure 4 | Page 7 |

### Supplementary Table 1. Performance comparison of the models for detecting CAD on external validation set.

| Model | AUC | 95% CI | P-value |
|---|---|---|---|
| AI imaging | 0.83 | 0.81-0.85 | Reference |
| Clinical Score | 0.80 | 0.77-0.82 | 0.02 |
| Ischemic TPD | 0.79 | 0.77-0.82 | <0.001 |
| Myocardial Flow Reserve | 0.75 | 0.72-0.78 | <0.001 |
| Stress MBF | 0.75 | 0.72-0.77 | <0.001 |
| Coronary Calcium | 0.69 | 0.66-0.72 | <0.001 |

Area under the receiver-operating characteristic curve (AUC) with 95% confidence interval (CI) for all artificial intelligence models and coronary calcium and ischemic total perfusion deficit (TPD) in all patients.

Abbreviations: AI – Artificial Intelligence; MBF – Myocardial Blood Flow.

### Supplementary Table 2. Sensitivity Comparison of AI and Quantitative Analysis in High-Risk Groups

| | n | AI | Ischemic TPD ≥ 5% |
|---|---|---|---|
| Left main stenosis ≥50% or ≥70% in other arteries | 674 | 566 | 530 |
| High risk groups | | | |
| Duke category 6 | 224 | 210 | 188 |
| Duke category 5 | 165 | 153 | 138 |
| | **n** | **AI** | **MFR < 2** |
| Left main stenosis ≥50% or ≥70% in other arteries | 674 | 602 | 514 |
| High risk groups | | | |
| Duke category 6 | 224 | 213 | 190 |
| Duke category 5 | 165 | 157 | 136 |
| | **n** | **AI** | **MBF < 1.8** |
| Left main stenosis ≥50% or ≥70% in other arteries | 674 | 607 | 540 |
| High risk groups | | | |
| Duke category 6 | 224 | 213 | 200 |
| Duke category 5 | 165 | 157 | 142 |

Comparison on the external testing set (n=1,278) with matched specificity. MFR and MBF represent the minimum values across the three vessels. AI model identified more true positives in each category over the quantitative analysis (e.g., ischemic TPD ≥5%). Duke category 6 is defined as left main ≥50% or 3-vessel ≥70%, or 2-vessel CAD including proximal LAD. Duke category 5 is defined as 2-vessel severe stenosis not including the proximal LAD, 1-vessel severe proximal LAD, or 3-vessel moderate stenosis ≥50%[1].

Abbreviations: AI – Artificial Intelligence; CAD – Coronary Artery Disease; LAD – Left Anterior Descending Artery; MBF – Myocardial Blood Flow; MFR – Myocardial Flow Reserve; TPD – Total Perfusion Deficit.

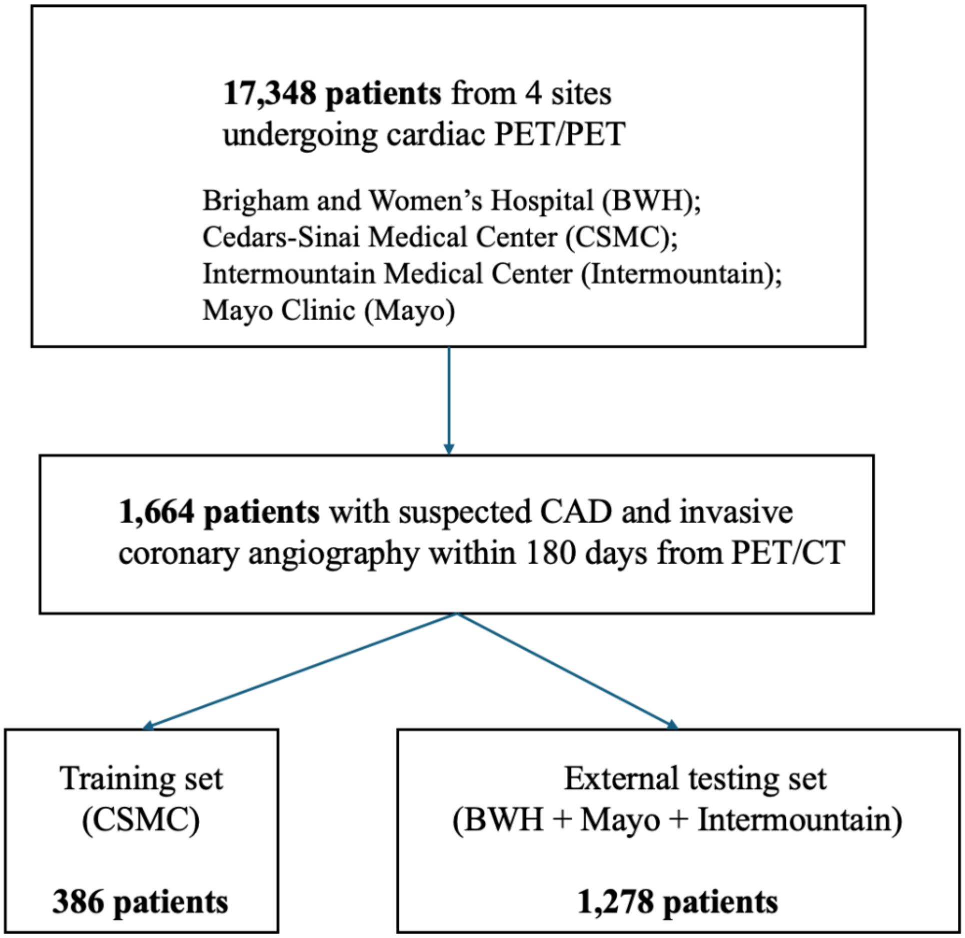

**Supplementary Figure 1. Flowchart of the study.** An illustration describes the inclusion criteria used in the analysis and how training set and external testing set were created for model development.

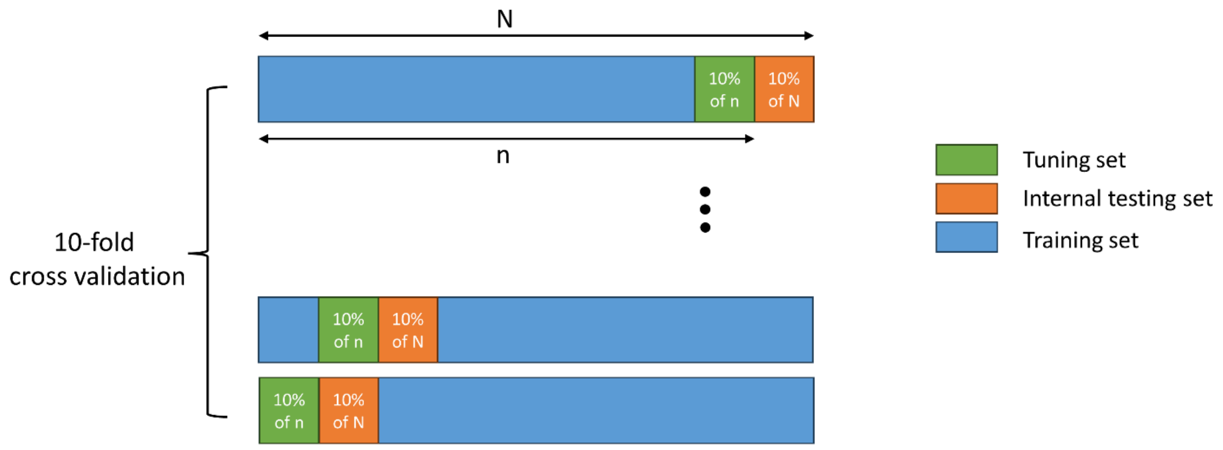

**Supplementary Figure 2. 10-Fold Cross-Validation for Training, Tuning, and Testing.** Model training is shown in blue, hyper-parameter tuning in green, and internal testing in orange. Hyper-parameters (e.g., the depth of the tree or the number of estimators) of XGBoost models are often empirically chosen and cannot be learned from model training. The recommended approach to discover the best values of hyper-parameters is by fitting the models with various combinations and assess their performance. To achieve this, a nested 10-fold cross-validation was implemented in each 10-fold subset (n): allocating 90% of this data for model fitting and 10% for tuning.

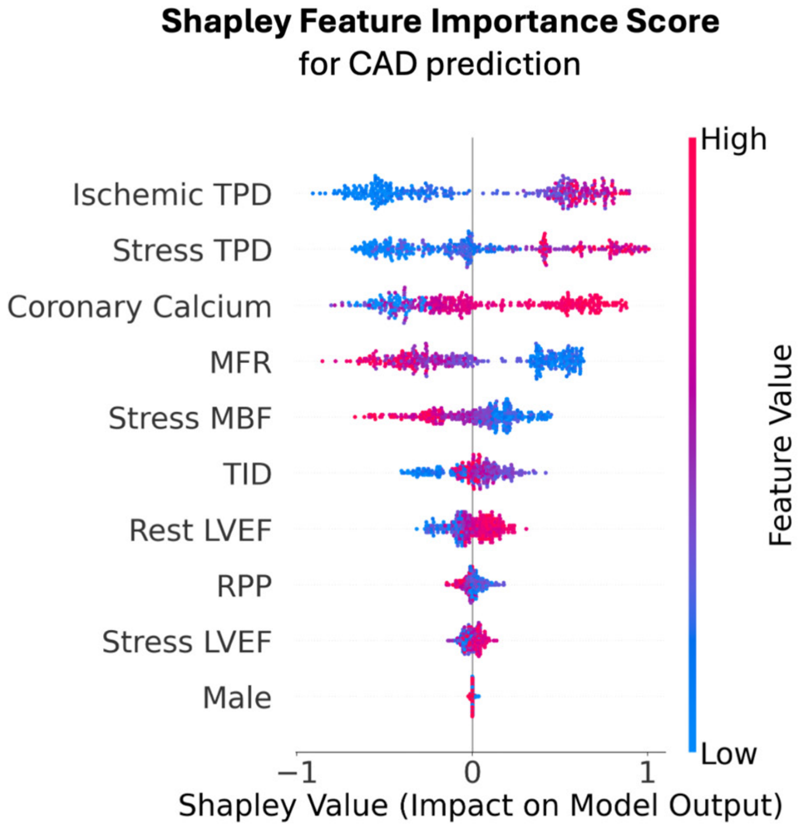

**Supplementary Figure 3.** Shapley feature importance for CAD prediction. Each dot represents an individual patient and a specific feature. The distance of each dot from the center line (0 on the x-axis) indicates how strongly that feature drives the prediction, either positively or negatively, for that patient. Abbreviations: LVEF – Left Ventricle Ejection Fraction; MBF – Myocardial Blood Flow; MFR – Myocardial Flow Reserve; RPP – Rate Pressure Product; TID – Transient Ischemic Dilation; TPD – Total Perfusion Deficit.

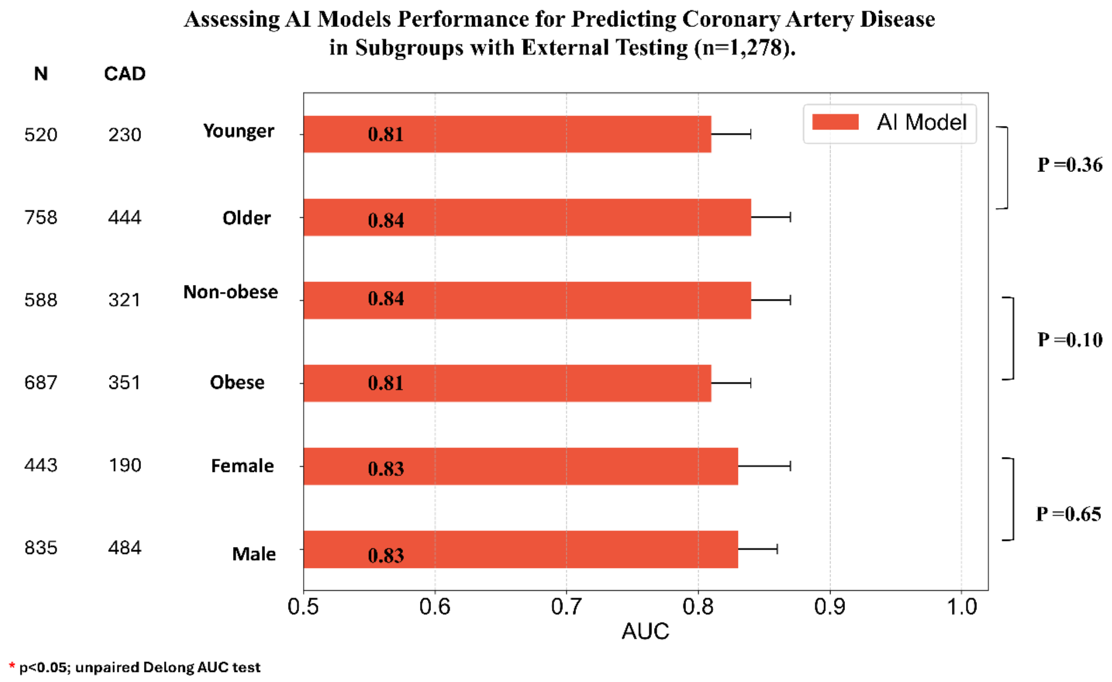

**Supplementary Figure 4. Performance of artificial intelligence (AI) for prediction of coronary artery disease (CAD) in subgroups.** AI models exhibit comparably robust performance in all subgroups. The younger group was defined as patients aged <65 years, while the older group was defined as patients aged ≥65 years. Similarly, the obese group was defined as patients with a BMI ≥30 kg/m², whereas the non-obese group was defined as patients with a BMI <30 kg/m². Race analysis was excluded due to the limited availability of data on minority race groups. P-values were generated using unpaired Delong test. Abbreviations: AUC – Area Under the Receiver Operating Characteristic Curve; BMI – Body Mass Index.

### References

1. Reynolds HR, Shaw LJ, Min JK, et al. Outcomes in the ISCHEMIA Trial Based on Coronary Artery Disease and Ischemia Severity. *Circulation* 2021; 144: 1024-1038. 20210909. DOI: 10.1161/CIRCULATIONAHA.120.049755.
